# Proteomic prediction of common and rare diseases

**DOI:** 10.1101/2023.07.18.23292811

**Authors:** Julia Carrasco-Zanini, Maik Pietzner, Jonathan Davitte, Praveen Surendran, Damien C. Croteau-Chonka, Chloe Robins, Ana Torralbo, Christopher Tomlinson, Natalie Fitzpatrick, Cai Ytsma, Tokuwa Kanno, Stephan Gade, Daniel Freitag, Frederik Ziebell, Spiros Denaxas, Joanna C. Betts, Nicholas J. Wareham, Harry Hemingway, Robert A. Scott, Claudia Langenberg

## Abstract

**Background:** For many diseases there are delays in diagnosis due to a lack of objective biomarkers for disease onset. Whether measuring thousands of proteins offers predictive information across a wide range of diseases is unknown.

**Methods:** In 41,931 individuals from the UK Biobank Pharma Proteomics Project (UKB-PPP), we integrated ∼3000 plasma proteins with clinical information to derive sparse prediction models for the 10-year incidence of 218 common and rare diseases (81 – 6038 cases). We compared prediction models based on proteins with a) basic clinical information alone, b) basic clinical information + 37 clinical biomarkers, and c) genome-wide polygenic risk scores.

**Results:** For 67 pathologically diverse diseases, a model including as few as 5 to 20 proteins was superior to clinical models (median delta C-index = 0.07; range = 0.02 – 0.31) and to clinical models with biomarkers for 52 diseases. In multiple myeloma, for example, a set of 5 proteins significantly improved prediction over basic clinical information (delta C-index = 0.25 (95% confidence interval 0.20 – 0.29)). At a 5% false positive rate (FPR), proteomic prediction (5 proteins) identified individuals at high risk of multiple myeloma (detection rate (DR) = 50%), non-Hodgkin lymphoma (DR = 55%) and motor neuron disease (DR = 29%). At a 20% FPR, proteomic prediction identified individuals at high-risk for pulmonary fibrosis (DR= 80%) and dilated cardiomyopathy (DR = 75%).

**Conclusions:** Sparse plasma protein signatures offer novel, clinically useful prediction of common and rare diseases, through disease-specific proteins and protein predictors shared across multiple diseases.

**(Funded by Medical Research Council, NIHR, Wellcome Trust.)**

## Introduction

A central challenge in precision medicine is the development of clinically useful tools for identifying individuals at high risk which may enable timely diagnosis, early initiation of treatment and improved patient outcomes^1^. Clinically recommended tools for predicting the risk of onset of diseases are widely used for heart attack and stroke (e.g., the AHA / ACC 10-year risk equation)^2^ but for very few other diseases. Across diverse diseases pathologies, diagnostic delays of months or years are reported from the initial onset of symptoms^3–5^. Over the last decades, single plasma proteins have become established as specific, diagnostic assays for a small number of diseases including BNP for heart failure, troponins for acute coronary syndromes and UCH-L1 and GFAP in traumatic brain injury^6^.

Plasma proteomics allows estimation of thousands of proteins and agnostic discovery studies not confined to a single disease of interest and represents a promising technology to accelerate progress towards this challenge. Plasma proteomic signatures capture health behaviours and current health status^7^, and may integrate the risk of “static” genetic^8,9^ and dynamic environmental determinants of disease. Translatable, parsimonious models have been described. For example, a sparse protein signature, containing as few as three proteins, improved identification of a high-risk group for diabetes which is currently missed by screening strategies.^10^

Whether plasma proteomics may offer clinically useful predictive or mechanistic information across a wide range of diseases, alone or in combination, is unknown for several reasons. First, previous proteomic studies have had too few participants to evaluate rare and common diseases. Secondly, previous studies of disease onset have focussed on a narrow set of common diseases^7,11–13^, rather than taking an agnostic discovery approach. Thirdly, previous studies have not reported screening metrics compared to clinical models (without proteins) which may inform integration into health records and translational evaluation.

We used data from the UK Biobank Pharma Proteomics Project (UKB-PPP), the largest proteomic experiment to date, to address the following objectives (i) to systematically interrogate the 10 year predictive potential of the measurable plasma proteome across 218 pathologically diverse diseases, over and above models based on information obtained in usual care (without and with clinical biomarkers) and polygenic risk scores (ii) to identify disease-specific protein predictors pointing to underlying aetiological mechanisms, compared to those shared across diseases (iii) to determine whether the screening metrics of proteomic signatures for diseases meet, or exceed, those for blood biomarkers used in current clinical practice.

## Methods

### Study design

We carried out a cohort study in the UKB-PPP to develop, validate and compare predictive models with and without proteins. UKB is a highly characterised longitudinal cohort of 500 000 adults. Individuals were excluded if they had missing data for age, sex and body mass index (BMI) or failed quality control (QC) criteria for proteomic measurements. The human biological samples were sourced ethically, and their research use was in accordance with the terms of the informed consent and under an IRB/EC approved protocol.

### Clinical risk information

Clinical risk information (without blood biomarkers) recommended as part of usual primary care, was obtained from UKB health questionnaires. This included: age at baseline, self-reported ethnicity, smoking status, alcohol consumption, paternal or maternal history for 15 individual diseases available (data-field IDs 20197 and 20110, **Table S1**), and measured BMI. For clinical risk information with blood biomarkers, we included 37 of the most widely performed blood tests (16 of these are based on proteins) which were assessed in all UKB participants (UKB Category 17518, 100081). Quality control of these ‘clinical biomarkers’ was done based on methods previously described^14,15^ and imputation was done using the missForest R package^16^ including additional information on age and sex.

### Proteomic profiling

Proteomic profiling was performed in EDTA-plasma samples from 54,893 UKB participants obtained at baseline as part of the UK Biobank Pharma Proteomics Project (UKB-PPP), using the Olink Explore 1536 and Expansion platforms, which captured 2923 unique proteins targeted by 2941 assays. Assay details^17,18^, sample selection and handling have been previously described^19^. The current study is based on participants from a randomly selected subset (N = 46,750). After quality control, we imputed missing NPX (normalised protein expression) values, using the missForest R package^16^, for all individuals who met the QC and inclusion criteria and had no more than 50% of missing values across all proteins, **Table S1-2, Supplementary Appendix**). Imputation was done per Olink panel, including additional information on age and sex.

### Incident disease definitions

We developed prediction models for 218 diseases, with more than 80 incident cases within 10 years of follow-up (censoring date was the 31^st^ of December 2020 or death date if this occurred first) in the random subset (N = 41,931, 193 diseases), or by including incident cases within the “consorTum-selected” subset (25 diseases) (**Table S1**). The 218 diseases include common and rare diseases, and diseases associated with high morbidity, high mortality, or both. Disease definitions were based on validated phenotypes described by Kuan *et al.*^20^ by integrating data from primary care, hospital episode statistics, cancer and death registries and from UKB health questionaries including self-reported illnesses. We excluded prevalent cases (first occurrence prior to or up to the baseline assessment visit) or incident cases recorded within the first 6 months of follow-up.

### Statistical analyses

We adapted a 3-step machine learning framework including (1) feature selection, (2) hyperparameter tuning and optimization, and (3) validation. Individuals were divided: 50% for feature selection, 25% for model optimization (training), and 25% for validation, for diseases with more than 800 cases; otherwise, into a 70% feature selection and model optimization set, and 30% for validation. Validation sets included non-overlapping individuals completely blinded to previous model development stages.

We performed feature selection among 2941 protein targets, or among the 37 clinical biomarkers by least absolute shrinkage and selection operator (LASSO) regression over 200 subsamples of the feature selection set. In each iteration, we ran 5-fold cross-validation over 3 repeats using a grid search to tune the hyperparameter lambda. We used the ROSE R package^21^ to address case imbalance. Selection scores were computed as the absolute sum of weights from the model with the optimal lambda from each of the 200 iterations and were used to identify the top 20 proteins or clinical biomarkers (**Supplementary Appendix**).

We used regularised cox regression to derive a “benchmark” clinical model, by 5-fold cross-validation in the optimisation or training set using the features described above. We tested improvement in models by adding onto the patient information: 1) 5 – 20 proteins, 2) 5 – 20 clinical biomarkers or 3) genome-wide polygenic scores^22^ (PGS, UKB category 301) (**Figure 1)**. For these comparisons, we trained and tested models including up to 5, 10 and 20 proteins or biomarkers and kept the best performing protein signature and biomarker signature. Validation was performed in the held-out test set, where we computed the concordance index (C-index) over 1000 bootstrap samples. Significant improvements between models were considered as those for which the 95 % confidence interval (95% CI) of the differences in the bootstrap C-index distributions did not include zero.

**Figure 1.**
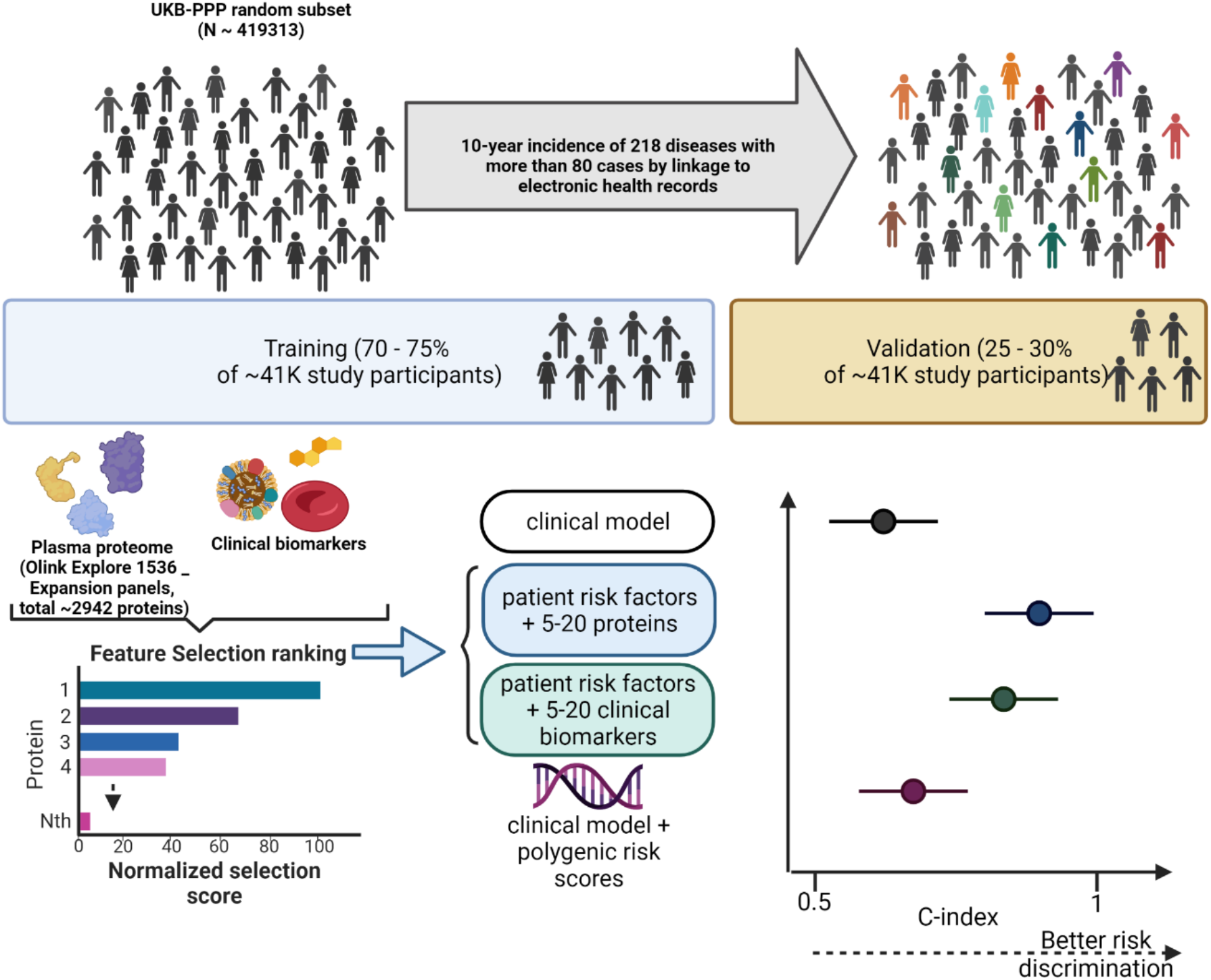
Study design. This cohort study is based on a random subset of UKB-PPP individuals (N = 41,931). All individuals were divided into training (including feature selection and optimisation steps) and validation sets to develop sparse protein-based predictors (including 5-20 proteins from the Olink Explore 1536 + Expansion panels) for 218 diseases defined using data from the UKB health-questionnaire, primary care, hospital episode statistics, cancer and death registries. Performance of protein-signatures was compared to clinical models, clinical biomarkers and genome-wide polygenic risk scores (PGS). Further details of methods are in the (Supplementary Appendix).

The screening metrics we calculated were: detection rates (DR) and likelihood ratios (LR) in the validation set at false positive rates (FPR) ranging from 5 to 40%. The FPR was calculated as FPR = false positives (FP) / (true negatives (TN)+ FP); and detection rates were calculated as DR = true positives (TP)/ (false negatives (FN) + TP). LRs were computed as LR = DR / FPR. All analyses were performed in R software version 4.1.1.

## Results

### Improvement in prediction by adding sparse protein signatures vs clinical biomarkers onto clinical models

Clinical models without blood biomarkers showed a median C-index = 0.64 (interquartile range = 0.58 – 0.72), achieving the highest performance for endocrine and cardiovascular diseases (**Figure S1, Table S3**). For 67 rare and common diseases (**Figure S2**), addition of 5 to 20 proteins significantly improved (95% confidence intervals of improvement in C-index > 0) clinical models (median increase in C-index = 0.07, range = 0.02 – 0.31) (**Figure 2a, Table S4**). Diseases for which proteins improved clinical models included multiple myeloma (delta C-index = 0.25 (95% confidence interval 0.20 – 0.29, LR = 6.55), non-Hodgkin lymphoma (delta C-index = 0.21 (0.14 – 0.28), LR = 6.08), pulmonary fibrosis (delta C-index = 0.09 (0.03 – 0.14), LR = 6.83), coeliac disease (delta C-index = 0.31 (0.21 – 0.38), LR=8.07), dilated cardiomyopathy (delta C-index = 0.17 (0.10 – 0.22), LR =6.97) and motor neuron disease (delta C-index = 0.11 (95% CI: 0.04 – 0.16), LR = 4.38) (**Figure 2a**). Across these 67 diseases, the median detection rate (at a 10% FPR, DR10) was 45.5% (range: 10.8 – 80.8 %), compared to 25% (range: 9.5 – 51.2%) for the clinical model (**Figure 2b, Table S5**). The median LR was 4.55 (range: 1.08 – 8.07) for these 67 diseases, representing improvements ranging from 0.12 – 6.92 over the clinical models (**Figure 2c**). For example, applying a protein-informed test for coeliac disease (LR = 8.08) would result in detecting 80.8% of cases, while retaining an acceptable proportion of 10% false-positives (**Figure S3**). Clinical models with blood biomarkers only significantly improved prediction over clinical models for 28 diseases (median delta C-index = 0.08, range = 0.01 – 0.28) (**Figure 3, Table S6**). For 52 of these diseases, protein-based models achieved higher LRs (range = 0.13 – 5.17) in comparison to clinical model with blood biomarkers (**Figure S4, Table S7**). Compared to the single most informative protein, sparse protein signatures (5-20 proteins) had an average 5.4% improvement in C-index over clinical models, across diseases that achieved significant improvements. For 64% of these, performance saturation was achieved by including a maximum of 5 to 10 proteins.

**Figure 2.**
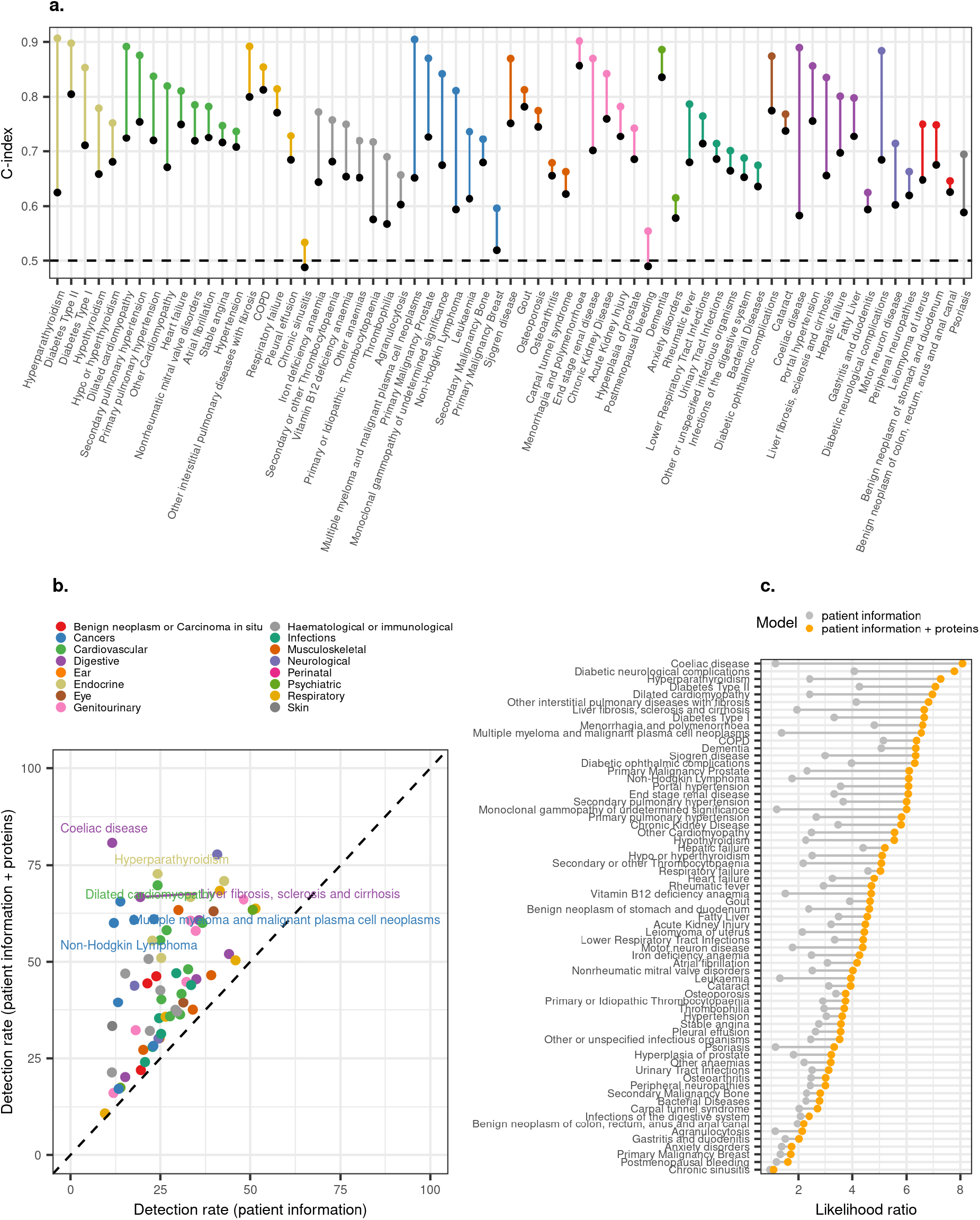
Improvement in predictive performance by addition of proteins onto basic patient risk factors for 67 incident diseases. **a,** Improvement in C-index by the addition of 5 – 20 proteins (coloured dots) over the benchmark patient-information model (black dots)**. b,** Comparison between detection rates (at a 10% false positive rate) achieved by protein-based and patient-information model**. c,** Improvement in likelihood ratios by the addition of 5 – 20 proteins (orange) over the benchmark clinical model (grey).

**Figure 3.**
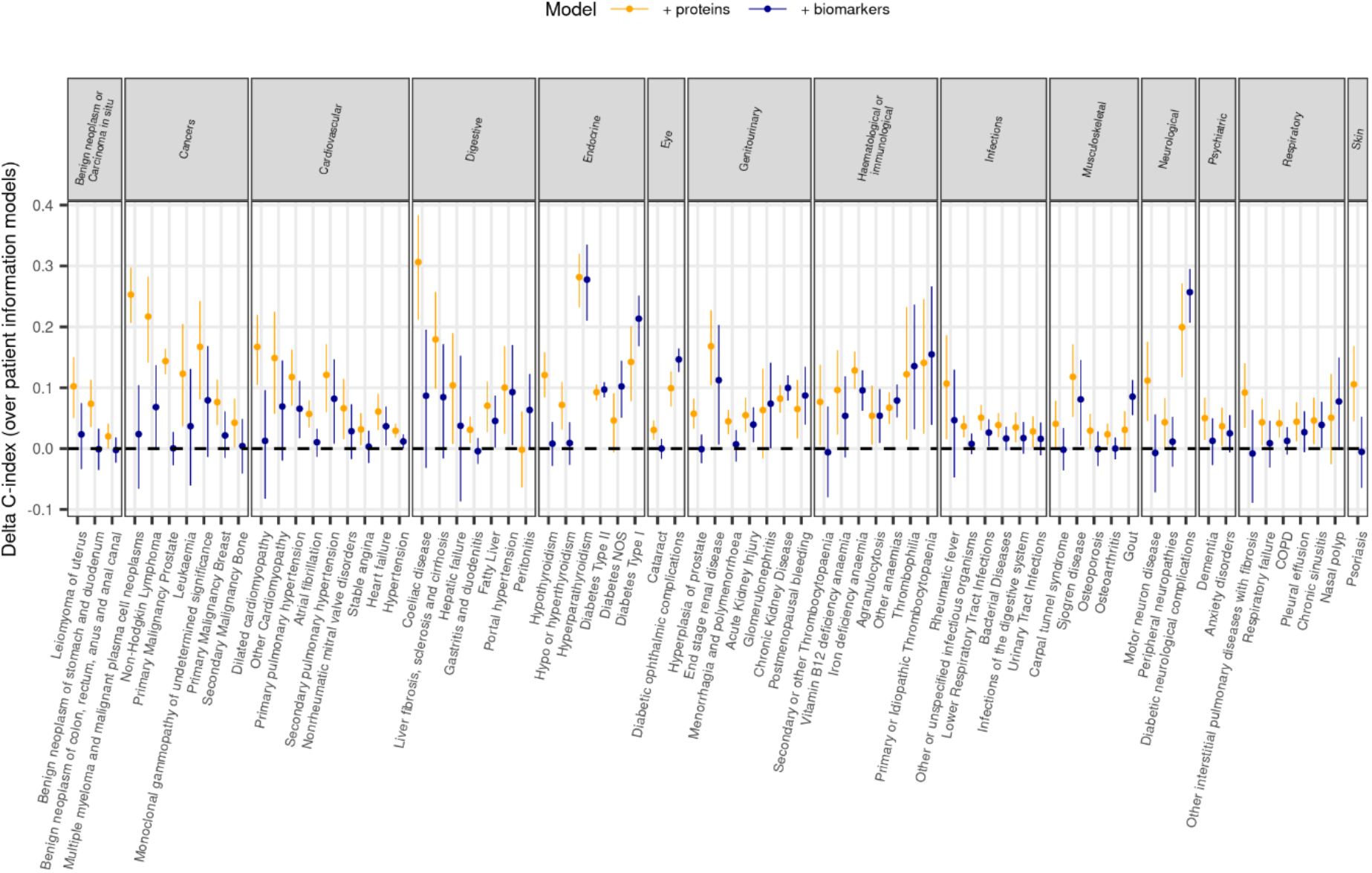
Comparison of predictive performance between proteins-based (patient information + proteins) and biomarker-based (patient information + biomarkers) models. **a,** Comparison of C-index by the addition of protein-based (coloured dots) or biomarker-based models (black dots) onto patient risk factors. We only show those diseases for which C-index was significantly improved by addition of either proteins or clinical biomarkers onto the patient risk factors.

### Proteins predicting multiple diseases

The 67 prediction models with clinically relevant improvements, included a total of 501 protein targets, of which 147 were selected for 2 or more (range: 2 - 16) diseases (**Figure S5**); most of which (∼89%) were selected across 2 or more clinical specialties (range: 2 - 9) (**Figure 4a**). On average, these had a relatively lower contribution for prediction of individual diseases, in comparison to highly specific proteins (**Figure 4b**). Age was the major correlate of 4 out of the 5 proteins that were predictive across more than 10 diseases and smoking status was the major correlate for CXCL17 (**Figure S6**), but these proteins still provided improvements in prediction over and above these conventional risk factors.

**Figure 4.**
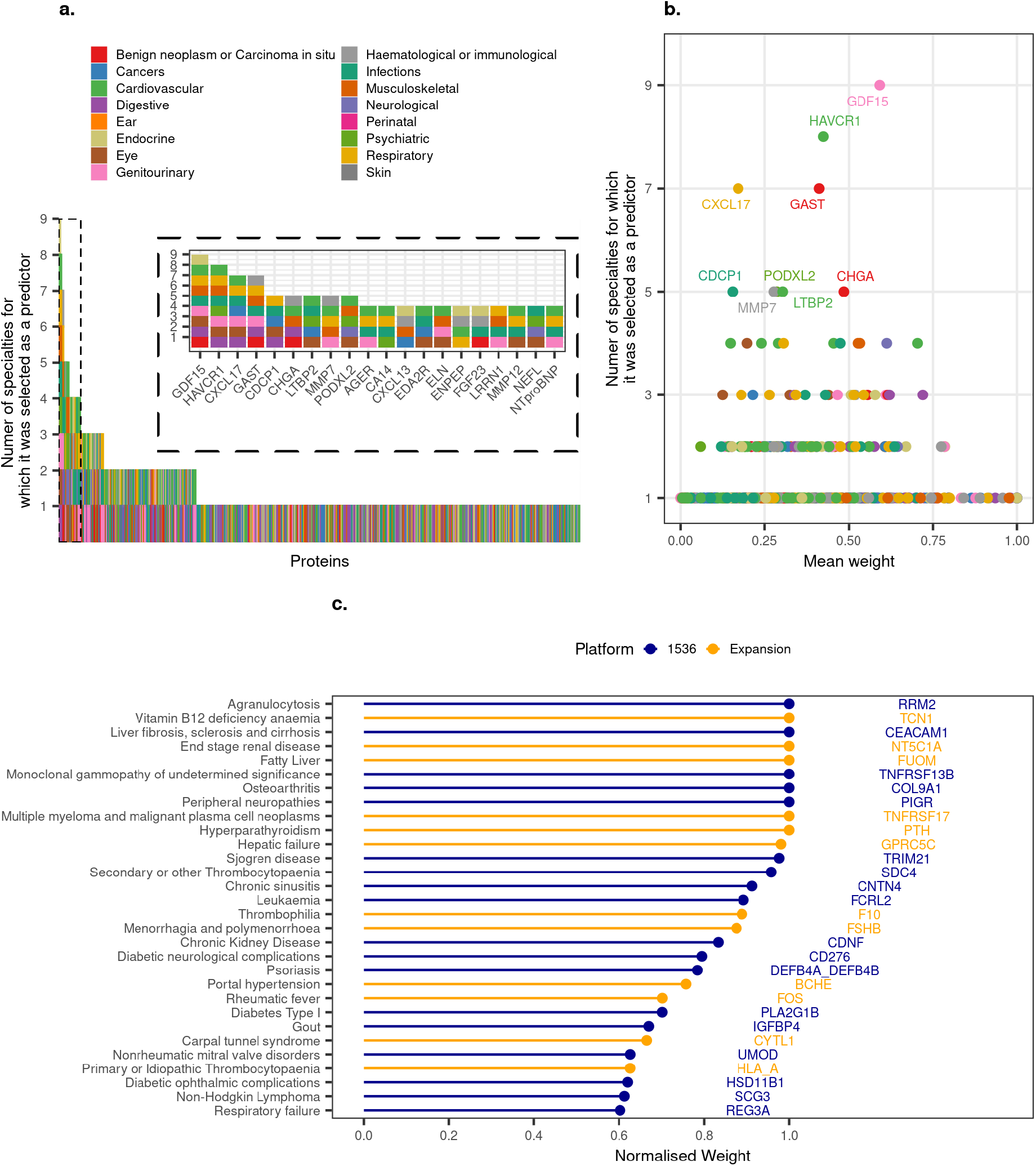
Disease specificity of predictor proteins. **a,** Number of disease groups for which a protein was selected as a predictor across the 88 diseases. These were diseases for which the C-index was significantly improved or improved by more than 0.4 over the patient information model. **b**, Average contribution of proteins across diseases. Average weights (normalised to the top predictor) from the optimised prediction models for each protein, across diseases for which it was selected as a predictor (out of the 88 improved diseases). **c,** Disease-specific proteins are shown as those selected for only one disease with a normalised weight > 0.6.

### Proteins specifically predicting one disease

We identified proteins solely and strongly predictive for only one disease (**Figure 4c, Table S8**), including TNF receptor superfamily member 17 (TNFRSF17 or B-Cell Maturation antigen), a specific predictor for multiple myeloma; and TNFRSF13B, a strong predictor of monoclonal gammopathy of undetermined significance (MGUS), a condition which precedes the development of multiple myeloma (at a rate of ∼1 in 100 MGUS cases developing multiple myeloma per year^23^). Here, we provide evidence that increased plasma levels of these receptors (**Table S9**) are strongly predictive of future onset for these blood cancers. Previous studies have already suggested an association between plasma TNFRSF17 and progression from MGUS to multiple myeloma^24^. Here we identified the added value of a 5-protein protein signature, which improved discrimination by 7% over patient risk factors + TNFRSF17 alone.

### PGS compared to clinical models and protein models

For 23 diseases for which PGS were available in UKB, we found that PGS significantly improved prediction over clinical models for only 7 diseases, but with clinically negligible improvements (median delta C-index = 0.03, range = 0.01 – 0.14) (**Table S10**). Proteins outperformed PGS for all of these, except for breast cancer (**Figure S7**).

### Screening metrics for protein and clinical models

We observed consistently superior screening metrics across all conditions for a wide range of FPRs (5%-40%; **Figure 5**). At a 20% FPR, proteomic prediction identified individuals at high-risk for pulmonary fibrosis (including CA4, CEACAM6, GDF15, SFTPD and WFDC2; DR=80%) and dilated cardiomyopathy (including HRC, TNNI3, TPBGL, NPPB, NTproBNP; DR=75%). At a low FPR (5%), proteomic prediction identified individuals at high risk for multiple myeloma (FCRLB, QPCT, SLAMF7, TNFRSF17, TNFSF13B; DR = 50%), non-Hodgkin lymphoma (BCL2, CXCL13, IL10, PDCD1, SCG3; DR = 55%) and motor neuron disease (including CST5, EGFLAM, NEFL, PODXL2 and TMED10; DR = 29%). In sensitivity analyses we found that adding a larger set of proteins included in Olink’s Explore Expansion panels (**Supplementary appendix**) did not generally improve model performance compared to the first release of 1463 proteins (**Figure S8, Table S4**). However, improvements for selected diseases were obtained by including a specific predictive biomarker (only captured in the Expansion panels), such as TCN1 (a vitamin B12 binding protein) for vitamin B12 deficiency anaemia, KLK3 (prostate-specific antigen) for prostate cancer or, F10 (a coagulation factor that converts prothrombin into thrombin) and PROS1 (an anticoagulant protein) for thrombophilia (**Figure S8**). Protein-based models trained on 10-year incidence performed equally well when restricting the follow-up time to 5 years (Pearson r = 0.96, **Figure S9a**), although patient information models appeared to have systematically lower performances indices up to 5-years (Pearson r = 0.88, **Figure S9b**).

**Figure 5.**
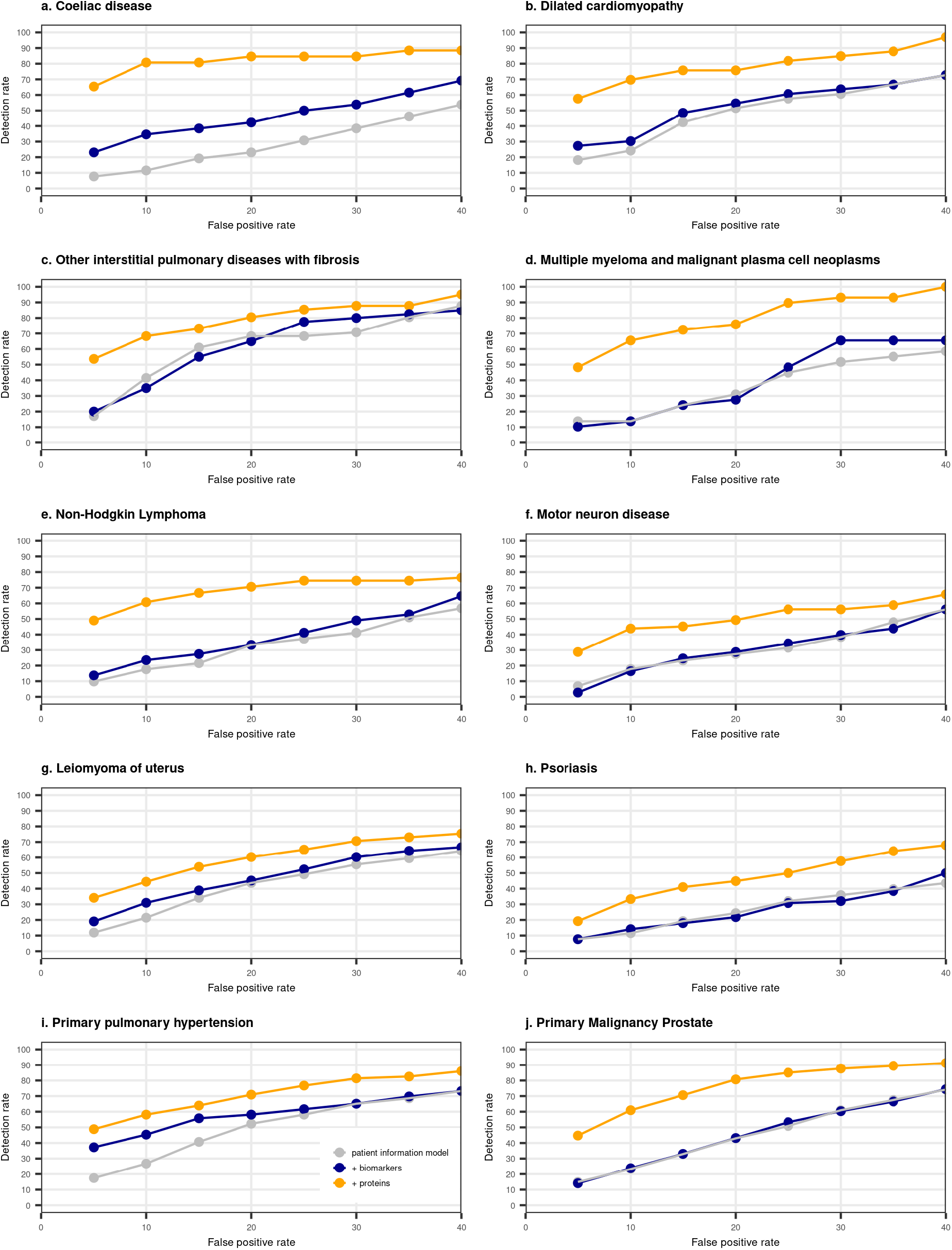
Detection rate curves. Detection rates across different false positive rate thresholds for selected examples identified as those most likely to benefit from proteomic prediction over patient risk factors, clinical biomarkers and PGS. Coeliac disease (TGM2, NOS2, ITGB7, CD160, PPP1R14D, RBP2, CCL25, MLN, FGF19, HMOX1, CEND1, MILR1, CDH2, CKMT1A_CKMT1B, CPA2, GTF2IRD1, SEPTIN3, BCL2L15, FABP2, HSD17B14). Dilated Cardiomyopathy (HRC, TNNI3, TPBGL, NPPB, NTproBNP). Other interstitial pulmonary disease with fibrosis (CA4, CEACAM6, GDF15, SFTPD and WFDC2). Multiple myeloma and malignant cell neoplasms (FCRLB, QPCT, SLAMF7, TNFRSF17, TNFSF13B). Non-Hodgkin Lymphoma (BCL2, CXCL13, IL10, PDCD1, SCG3). Motor neuron disease (CST5, EGFLAM, NEFL, PODXL2 and TMED10). Leiomyoma of uterus (BMP4, CDH3, CHRDL2, DNPEP, FGF23, GFRAL, LEFTY2, PAEP, SEZ6L2, TSPAN1). Psoriasis (DEFB4A_DEFB4B, IL19, KCTD5, PI3, PRKD2). Primary pulmonary hypertension (NPPB, NTproBNP, ROBO2, ENPEP, FGFBP2, LTBP2, SFRP1, ACP5, SPON1, CA4, SLC34A3, ACE2, AHSG, SERPINA7, SLC44A4, CDC123, SPINK8, LYPLA2, S100A3, MFAP4). Primary Malignancy Prostate (ADAMTS15, IL17A, INSL3, KLK3, LECT2, LTBP2, PRR5, SCARF2, SPINT3, TSPAN1).

## Discussion

We demonstrate the potential of sparse protein signatures to improve the prediction of disease onset across common and rare diseases. By integrating ∼3000 broad capture plasma proteins with EHRs, we showed that for 52 of 218 diseases studied, adding proteins was the single best prediction model, not only superior to commonly used patient characteristics, but also to a large array of biomarkers in clinical use and PGS (where available). Broad-capture proteomic technologies offer for many diseases new possibilities to address delays in diagnosis, the first blood-based biomarkers and the first evidence of clinically useful prediction models compared to current practice (**Table S11**). Plasma proteomic signatures may inform the need for, and design of, therapeutic clinical trials.

The wide spectrum of diseases that we studied enabled discovery of disease-proteomic signatures with the strongest screening metrics. The proteomic signatures that we report have screening metrics which were comparable to, or exceeded, those of blood tests currently used as diagnostic tests (for other diseases). Previous studies in a small number of diseases have investigated the predictive^7,11–13^ or prognostic^25^ potential of the circulating proteome. We found that for almost two-thirds (61%) of the superior protein models, a positive test, i.e., a predicted risk above the risk cut-off, translated into a four-fold increased risk of developing the disease compared to a negative one. Specifically, for 14 diseases, the LR achieved by protein-based models was higher than for a signature including prostate specific antigen (KLK3) for prostate cancer, which is used in currently implemented screening programs^26^. Sparse protein signatures (5-20 proteins) offer the opportunity to assess a limited set of proteins at a cost much below a discovery proteomic assay. The fact, that we identified strong predictive signatures in the non-fasting UKB samples further suggested feasibility of measurement in clinical practice.

We identified specific and strongly predictive proteins, pointing to underlying pathways conferring disease risk. Here we show that up to 10 years prior to diagnosis, higher plasma levels of TNFRSF17 and TNFRSF13B, receptors for BAFF and APRIL, were strong, specific predictors of increased risk of multiple myeloma and MGUS, respectively. These signalling pathways have been shown to promote multiple myeloma growth^27,28^. In turn, decreased plasma TNFSF13B, was further shown to be predictive of higher risk for multiple myeloma. Anti-TNFRSF17 agents, including antibody-drug conjugates (ADCs), T-cell engagers bispecific antibodies and cellular therapy with chimeric antigen receptor T cells (CAR-T), are approved for the treatment of refractory multiple myeloma^29–33^. Clinical trials exploring earlier implementation have started providing evidence for the safety and effectiveness of CAR-T cells in early lines of treatment^34^. Our results demonstrated the potential for implementation of proteomic screening, in a preventative manner even years before the onset of overt multiple myeloma, to identify the subgroup of individuals at highest risk, and highlight the possibility to test whether they represent those who would eventually benefit the most from anti-TNFRSF17 as earlier lines of treatment. Pulmonary fibrosis may be delayed due to misdiagnosis of other common respiratory or cardiovascular diseases^35^. The proteomic signature should be evaluated to identify who might benefit from enhanced surveillance through lung function tests and lung imaging, potentially enabling early treatment to maximise preservation of lung function^36^. For dilated cardiomyopathy, proteomic signatures could be evaluated for their potential to inform ECG and echo surveillance in people without a known genetic cause (up to 60% of cases^37,38^).

We found proteins predictive across multiple diseases and clinical specialties, consistent with shared aetiologies, including adaptations to ageing. Gastrin, for example, is well known for its role in production of hydrochloric acid, gastric motility and associations with gastrointestinal cancers and digestive system diseases^39^. However, our results highlighted associations with a wider range of diseases, including vitamin deficiencies, osteoporosis, infections and acute kidney injury. Proof-of-principle studies suggested that a single “omics” domain may predict risk of onset across multiple diseases^40^. Therefore, our results point to the potential for leveraging pleiotropic proteins to develop a customized, small signature for prediction across multiple diseases.

Our study has important limitations. Firstly, our results require validation in external studies, in ethnically diverse populations and in cohorts with differing pre-test probabilities of disease (UKB has a healthy participant effect^41^). Secondly, although we report the largest proteomic experiment to date, larger sample sizes are required to estimate detection rates for rarer diseases, and over shorter clinically relevant time frames (e.g., 1-5 years). Thirdly, evaluations against clinical diagnostic markers not available in UK Biobank are required including M-protein for multiple myeloma, and IgA/ IgG antibodies and anti-transglutaminase for coeliac disease. Fourthly, clinical translation will require development and validation of absolute quantification protein assays as opposed to the relative quantification provided by current proteomic platforms.

In conclusion, we demonstrated that sparse plasma protein signatures when integrated with electronic health records may offer novel, clinically useful prediction of common and rare diseases, through disease-specific proteins and protein predictors shared across multiple diseases.

## Data Availability

All individual level data is publicly available to bona fide researchers from the UK Biobank (https://www.ukbiobank.ac.uk/).

## Acknowledgements

All UK Biobank data was accessed in accordance with GlaxoSmithKline’s UK Biobank Application #20361 and the UKB-PPP Consortium Application #65851. We would like to acknowledge the UK Biobank participants for their dedication to participating in ongoing research and electronic health record linkage. This work and the incredible work of other UK Biobank researchers would not have been possible without their dedication to science.

## References

1. Bobrowska A, Murton M, Seedat F, et al. Targeted screening in the UK: A narrow concept with broad application. Lancet Reg Health Eur 2022;16:100353.

2. Goff DC, Jr., Lloyd-Jones DM, Bennett G, et al. 2013 ACC/AHA guideline on the assessment of cardiovascular risk: a report of the American College of Cardiology/American Heart Association Task Force on Practice Guidelines. Circulation 2014;129:S49–73.

3. Koshiaris C, Oke J, Abel L, Nicholson BD, Ramasamy K, Van den Bruel A. Quantifying intervals to diagnosis in myeloma: a systematic review and meta-analysis. BMJ Open 2018;8:e019758.

4. Hoyer N, Prior TS, Bendstrup E, Shaker SB. Diagnostic delay in IPF impacts progression-free survival, quality of life and hospitalisation rates. BMJ Open Respir Res 2022;9.

5. Abo-Tabik M, Parisi R, Morgan C, et al. Mapping opportunities for the earlier diagnosis of psoriasis in primary care settings in the UK: results from two matched case-control studies. Br J Gen Pract 2022;72:e834–e41.

6. Helmrich I, Czeiter E, Amrein K, et al. Incremental prognostic value of acute serum biomarkers for functional outcome after traumatic brain injury (CENTER-TBI): an observational cohort study. Lancet Neurol 2022;21:792–802.

7. Williams SA, Kivimaki M, Langenberg C, et al. Plasma protein patterns as comprehensive indicators of health. Nat Med 2019;25:1851–7.

8. Torkamani A, Wineinger NE, Topol EJ. The personal and clinical utility of polygenic risk scores. Nat Rev Genet 2018;19:581–90.

9. Polygenic Risk Score Task Force of the International Common Disease A. Responsible use of polygenic risk scores in the clinic: potential benefits, risks and gaps. Nat Med 2021;27:1876–84.

10. Carrasco-Zanini J, Pietzner M, Lindbohm JV, et al. Proteomic signatures for identification of impaired glucose tolerance. Nat Med 2022;28:2293–300.

11. Gadd DA, Hillary RF, Kuncheva Z, et al. Blood protein levels predict leading incident diseases and mortality in UK Biobank. medRxiv 2023:2023.05.01.23288879.

12. Ho JE, Lyass A, Courchesne P, et al. Protein Biomarkers of Cardiovascular Disease and Mortality in the Community. J Am Heart Assoc 2018;7.

13. Williams SA, Ostroff R, Hinterberg MA, et al. A proteomic surrogate for cardiovascular outcomes that is sensitive to multiple mechanisms of change in risk. Sci Transl Med 2022;14:eabj9625.

14. Sinnott-Armstrong N, Tanigawa Y, Amar D, et al. Genetics of 35 blood and urine biomarkers in the UK Biobank. Nat Genet 2021;53:185–94.

15. Vuckovic D, Bao EL, Akbari P, et al. The Polygenic and Monogenic Basis of Blood Traits and Diseases. Cell 2020;182:1214–31 e11.

16. Stekhoven DJ, Buhlmann P. MissForest--non-parametric missing value imputation for mixed-type data. Bioinformatics 2012;28:112–8.

17. Wik L, Nordberg N, Broberg J, et al. Proximity Extension Assay in Combination with Next-Generation Sequencing for High-throughput Proteome-wide Analysis. Mol Cell Proteomics 2021;20:100168.

18. Zhong W, Edfors F, Gummesson A, Bergstrom G, Fagerberg L, Uhlen M. Next generation plasma proteome profiling to monitor health and disease. Nat Commun 2021;12:2493.

19. Sun BB, Chiou J, Traylor M, et al. Genetic regulation of the human plasma proteome in 54,306 UK Biobank participants. bioRxiv 2022:2022.06.17.496443.

20. Kuan V, Denaxas S, Gonzalez-Izquierdo A, et al. A chronological map of 308 physical and mental health conditions from 4 million individuals in the English National Health Service. Lancet Digit Health 2019;1:e63–e77.

21. Nicola Lunardon, Giovanna Menardi, Torelli N. ROSE: a Package for Binary Imbalanced Learning. The R Journal 2014;6:78–9.

22. Thompson DJ, Wells D, Selzam S, et al. UK Biobank release and systematic evaluation of optimised polygenic risk scores for 53 diseases and quantitative traits. medRxiv 2022:2022.06.16.22276246.

23. Zingone A, Kuehl WM. Pathogenesis of monoclonal gammopathy of undetermined significance and progression to multiple myeloma. Semin Hematol 2011;48:4–12.

24. Visram A, Soof C, Rajkumar SV, et al. Serum BCMA levels predict outcomes in MGUS and smoldering myeloma patients. Blood Cancer J 2021;11:120.

25. Ganz P, Heidecker B, Hveem K, et al. Development and Validation of a Protein-Based Risk Score for Cardiovascular Outcomes Among Patients With Stable Coronary Heart Disease. JAMA 2016;315:2532–41.

26. Pinsky PF, Parnes H. Screening for Prostate Cancer. New England Journal of Medicine 2023;388:1405–14.

27. Tai YT, Acharya C, An G, et al. APRIL and BCMA promote human multiple myeloma growth and immunosuppression in the bone marrow microenvironment. Blood 2016;127:3225–36.

28. Shen X, Guo Y, Qi J, Shi W, Wu X, Ju S. Binding of B-cell maturation antigen to B-cell activating factor induces survival of multiple myeloma cells by activating Akt and JNK signaling pathways. Cell Biochem Funct 2016;34:104–10.

29. van de Donk N, Usmani SZ, Yong K. CAR T-cell therapy for multiple myeloma: state of the art and prospects. Lancet Haematol 2021;8:e446–e61.

30. Moreau P, Garfall AL, van de Donk N, et al. Teclistamab in Relapsed or Refractory Multiple Myeloma. N Engl J Med 2022;387:495–505.

31. Raje N, Berdeja J, Lin Y, et al. Anti-BCMA CAR T-Cell Therapy bb2121 in Relapsed or Refractory Multiple Myeloma. N Engl J Med 2019;380:1726–37.

32. Mikkilineni L, Kochenderfer JN. CAR T cell therapies for patients with multiple myeloma. Nat Rev Clin Oncol 2021;18:71–84.

33. Sammartano V, Franceschini M, Fredducci S, et al. Anti-BCMA novel therapies for multiple myeloma. Cancer Drug Resist 2023;6:169–81.

34. Garfall AL, Cohen AD, Susanibar-Adaniya SP, et al. Anti-BCMA/CD19 CAR T Cells with Early Immunomodulatory Maintenance for Multiple Myeloma Responding to Initial or Later-Line Therapy. Blood Cancer Discov 2023;4:118–33.

35. Guenther A, Krauss E, Tello S, et al. The European IPF registry (eurIPFreg): baseline characteristics and survival of patients with idiopathic pulmonary fibrosis. Respir Res 2018;19:141.

36. Maher TM, Strek ME. Antifibrotic therapy for idiopathic pulmonary fibrosis: time to treat. Respir Res 2019;20:205.

37. Harakalova M, Kummeling G, Sammani A, et al. A systematic analysis of genetic dilated cardiomyopathy reveals numerous ubiquitously expressed and muscle-specific genes. Eur J Heart Fail 2015;17:484–93.

38. Sweet M, Taylor MR, Mestroni L. Diagnosis, prevalence, and screening of familial dilated cardiomyopathy. Expert Opin Orphan Drugs 2015;3:869–76.

39. Duan S, Rico K, Merchant JL. Gastrin: From Physiology to Gastrointestinal Malignancies. Function (Oxf) 2022;3:zqab062.

40. Buergel T, Steinfeldt J, Ruyoga G, et al. Metabolomic profiles predict individual multidisease outcomes. Nat Med 2022;28:2309–20.

41. Fry A, Littlejohns TJ, Sudlow C, et al. Comparison of Sociodemographic and Health-Related Characteristics of UK Biobank Participants With Those of the General Population. Am J Epidemiol 2017;186:1026–34.

