## Supplemental Methods and Figures for "Proteomic prediction of common and rare diseases"

**Supplementary Appendix**

### Supplementary methods

#### *Study design*

The UK Biobank (UKB) study is a population-based cohort of around half a million participants from the United Kingdom aged between 40 and 59 years who were recruited between 2006 and 2010 (baseline assessment). Deep phenotype and genetic data are available for participants, including blood and urine biomarkers, whole-body imaging, lifestyle indicators, physical and anthropometric measurements, genome-wide genotyping, exome and genome sequencing. Follow-up is currently ongoing, and participants are further linked to routinely collected electronic health records. Detailed information is available at https://biobank.ndph.ox.ac.uk/showcase/.

Proteomic profiling was performed in EDTA-plasma samples from 54,893 UKB participants as part of the UK Biobank Pharma Proteomics Project (UK-PPP). Details of the sample selection and sample handling have been previously described[1]. Briefly, the study design included three elements: (1) a randomised subset of 46,750 individuals, (2) 6,856 individuals selected by the UKB-PPP consortium members ("consortium selected"), in which proteomic profiling was done on samples from the baseline assessment, and (3) 1,268 individuals who participated in a COVID-19 imaging study with repeated imaging at multiple visits. While the randomised subset was representative of the entire UKB population, "consortium selected" participants had different baseline characteristics for common risk factors (on average older, higher BMI and more smokers) and were enriched in cases for 122 different diseases[1]. Therefore, we based analyses on the randomised subset, with the exception of 25 less frequent diseases for which we included additional incident cases occurring within the "consortium-selected" participants.

#### *Proteomic data processing and quality control*

Proteomic profiling was done using the Olink Explore 1536 and Expansion platforms, which captured 2923 unique proteins targeted by 2941 assays. Assay details have been previously described[2,3]. Briefly, OlinkÒ relies on proximity extension assays, which targets proteins by pairs of antibodies conjugated to complimentary oligonucleotides. Upon binding to their target protein, hybridization between probes enables amplification and subsequent relative quantification through next generation sequencing. Protein targeting assays are grouped across four 384-plex panels: Inflammation, oncology, Cardiometabolic and Neurology. Olink's internal controls involve an incubation (a non-human antigen with matching antibodies), extension (IgG conjugated with a matching oligo pair) and amplification controls (synthetic double stranded DNA). Additional external controls are included in each plate, namely negative, plate and sample controls. Limit of detection values are calculated for each protein targeting assay per plate based on negative controls run in triplicate. Normalised protein expression

(NPX) values are generated by normalisation to the extension control, log2 transformation and further normalisation to the plate controls. Samples are flagged with a warning if NPX values from internal controls are not within ± 0.3 NPX from the plate median across an abundance block, or if the mean assay count for a sample is less than 500. Assays are flagged with a warning if the median from the negative control triplicated are deviate more than 5 standard deviations (SD) from predefined values set by Olink. We excluded (1) participants that were removed from the study and (2) samples that were defined as outliers. Outliers included individuals for which standardised first or second principal component values were further than 5 SD from the mean; or had a median NPX or interquartile range (IQR) of NPX greater than 5SD for the mean median or mean IQR. Individual data points with sample or assay warnings, or those belonging to 70 plates which failed to satisfy QC criteria were set to missing.

***Biomarker data and quality control***

We used data on 28 blood biomarkers (UKB Category 17518) and 9 blood cell traits (UKB Category 100081) (leukocyte, lymphocyte, monocyte, neutrophil, eosinophil, basophil, platelet count, haemoglobin concentration and haematocrit percentage), and refer to these 37 blood-based tests[4] (**Table S12**) as clinical biomarkers. Oestrogen and rheumatoid factor were not included in the analyses given these had more than 50% of missing values.  For the n=9 blood cell traits, we excluded blood-cell measures from individuals with extreme values or relevant medical conditions as described previously[5]. Relevant medical conditions for exclusion included pregnancy at the time the complete blood count was performed, congenital or hereditary anaemia, HIV, end-stage kidney disease, cirrhosis, blood cancer, bone marrow transplant, and splenectomy. Extreme measures were defined as leukocyte count >200x10*9*/L or >100x10*9*/L with 5% immature reticulocytes, hemoglobin concentration >20 g/dL, hematocrit >60%, and platelet count >1000x10*9*/L.

***Identification of best performing sparse protein and biomarker signature***

The 20 proteins with the highest feature selection scores were taken forward for optimisation of a model including the patient risk factors + 20 proteins, using regularised cox regression by 5-fold cross-validation (optimisation set, or feature selection set for diseases with less than 800 cases). To further identify sparser predictor sets, the top 5 and top 10 proteins were identified as those with the highest product of the weights from optimised models (patient risk factors + top 20 proteins) and feature selection scores. Optimisation of patient information + 5 or 10 proteins was similarly done by regularised cox regression by 5-fold cross-validation (optimisation set) and performance was tested in the validation set, by computing the C-index over 1000 bootstrap samples. Finally, models based on the top 5 proteins alone (without any patient risk factors) were further trained and tested in the same manner. The protein-based model (5-proteins alone, patient information + 5 proteins, patient information + 10 proteins or patient information + 20 proteins) which achieved the highest improvement in C-index over the patient-information model was kept as the final model. In a similar manner we identified the best set of 5, 10 or biomarker-based model which was kept as the final model for each of the diseases.

***Performance of prediction models for 5-year incidence***

The performance of patient information and patient information + protein models trained to predict the risk of 10-year incidence, was tested for 5-year incidence (same validation sets). This was tested for diseases for which 10-year incidence prediction (C-index) was significantly improved or improved by more than 4%, and had at least 20 incident cases within 5 years of follow-up in the validation set (54 diseases).

***Predictive performance of plasma proteins capture in the Olink 1536 panels vs Olink 1536 + Expansion panels***

We further repeated the entire procedure (that is feature selection, model optimisation and testing) on the first subset of Olink Explore 1536 proteins, using the exact same data splits for comparability (that is, the same individuals used in this analysis as those used in training/testing for the main analyses done on 1536 + Expansion proteins). Given we observed that for few selected examples, models based on the first set of ~1.5K proteins achieved higher performance, we compared stability of the feature selection procedures in both analyses.

***Proportion of variance explained in protein plasma levels***

We used the variancePartition R package[6] to estimate the proportion of variance explained in plasma levels of each of the proteins by a joint model including age, sex, body mass index, smoking status and the Elixhauser Comorbidity index[7] as explanatory variables. Briefly, this method fits a linear mixed model and estimates the proportion of variance explained attributed to each of the explanatory variables. We used this framework to identify the major correlates for each of the 5 proteins. We compared the proportion of variance explained by each of the variables for these 5 proteins, to the average proportion of variance explained across all other proteins.

**Supplementary results**

As a sensitivity analysis, we tested all optimised models in individuals from the validation set that had no missing values (for the proteins from the final model) to assess the quality of the imputation procedure. We observed good agreement between performance metrics derived in the test set which included a small proportion of imputed protein values and those derived from individuals with no missing data (Pearson r = 0.94).

We further performed an additional sensitivity analysis for 19 of the 25 diseases, for which incident cases among consortium selected participants were included. For these 19 diseases, there were at least 60 incident cases within the random subset of UKB-PPP, enabling demonstrating good agreement in predictive performance from the main analyses and by excluding consortium-selected incident cases from the test set (Pearson r = 0.97). This demonstrated no strong bias introduced from inclusion of participants who were selected based on specific characteristics or genetic risk of specific diseases.

We performed an additional analysis, to rule out the possibility that a statistical artifact could lead to the observed inverse relationship between incident case numbers and the improvement in C-index achieved proteins. We used hypertension (the disease with the highest number of incident cases) as an example to run this sensitivity analysis, in which we restricted selection of the number of incident cases to 80, 100, 150, 250, 500, 1000, 2000. We repeated the entire framework, including, feature selection, model optimization and validation, in these different configurations including fewer incident cases. We showed there was no inflation in the improvements in C-index achieved by adding proteins onto the patient information model, when restricting the analyses to fewer incident cases (**Table S13**).

### Supplementary figures

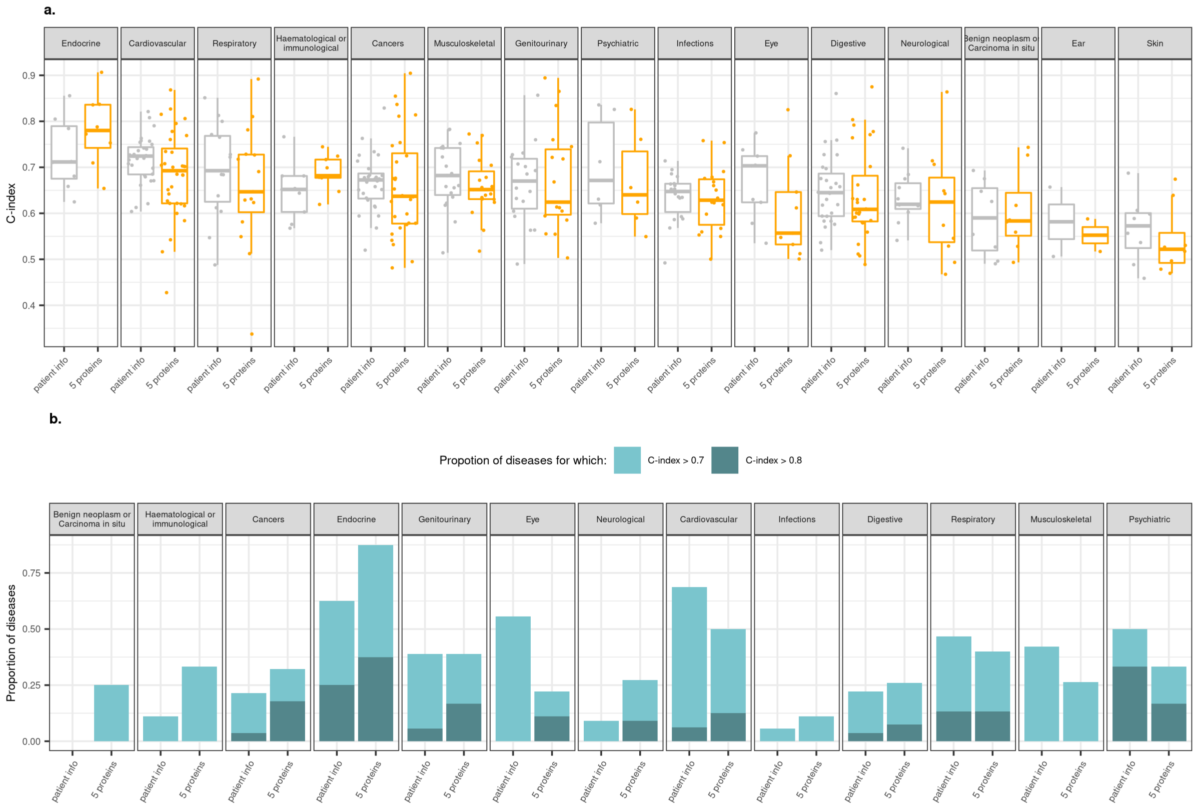

**Figure S1. Predictive performance of 5 proteins alone compared to the benchmark patient derived information models in the test set. a,** Distribution of the predictive performance (C-index) achieved by 5 proteins (orange) and patient risk factors (grey) within each disease category. **b,** Proportion of diseases within each category for which the patient risk factors or the top 5 proteins achieved a C-index > 0.7 (full bar height). Bar heights coloured in darker blue represent the proportion of diseases for which the C-index >0.8.

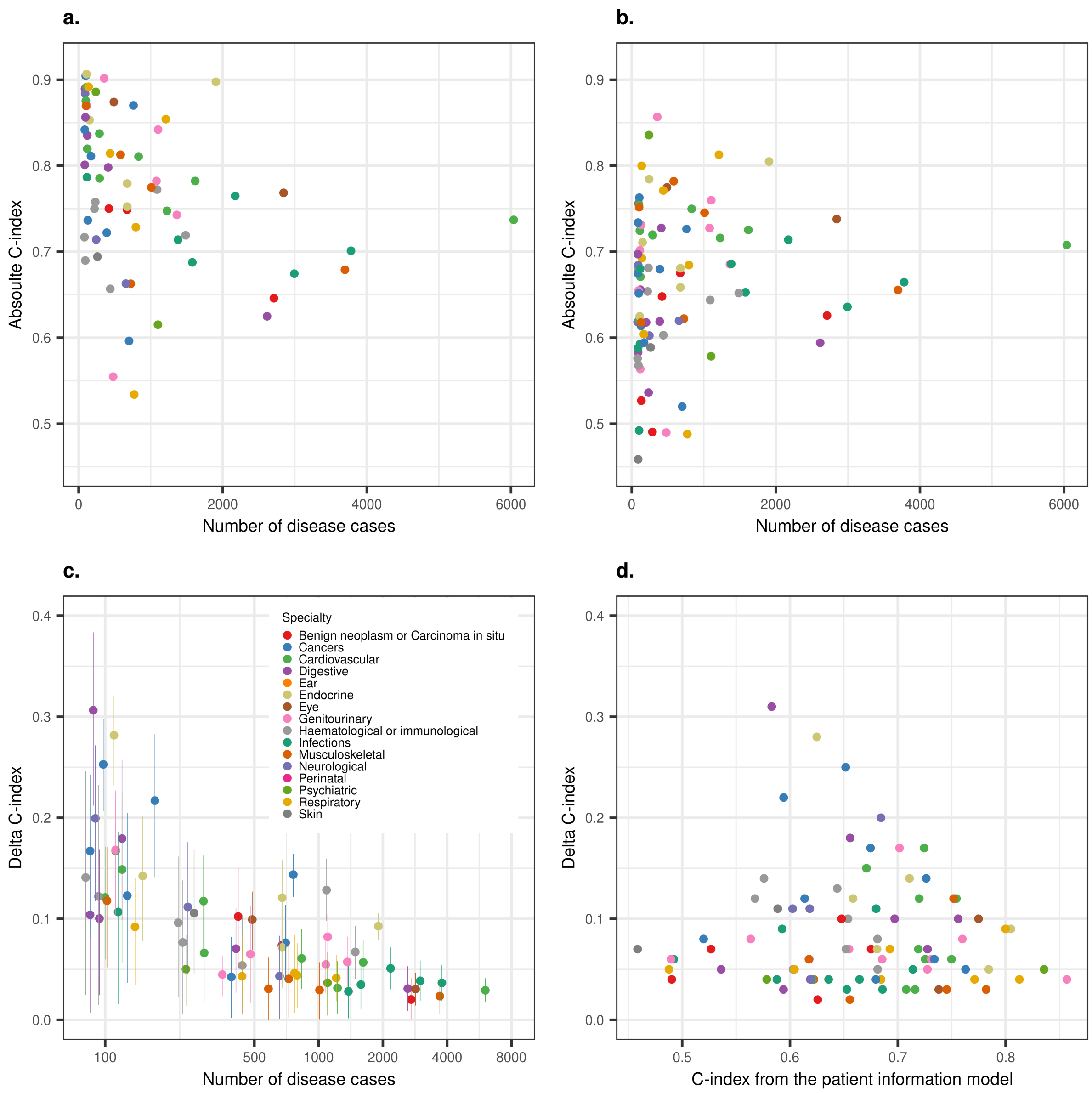

**Figure S2. Predictive performance is not related with the number of incident cases. a,** Predictive performance (C-index) of protein-based models, across 88 diseases for which these outperformed patient information models (or improved performance by 4%), was not correlated with the number of incident cases within 10 years of follow-up. **b,** Predictive performance (C-index) of patient information models was not correlated with the number of incident cases within 10 years of follow-up. **c,** Improvement in predictive performance (delta C-index) of protein-based models over patient information models appeared to be the largest for diseases less frequent among the UKB population. **d,** Improvement in predictive performance (delta C-index) of protein-based models was not correlated with baseline prediction of the patient information models.

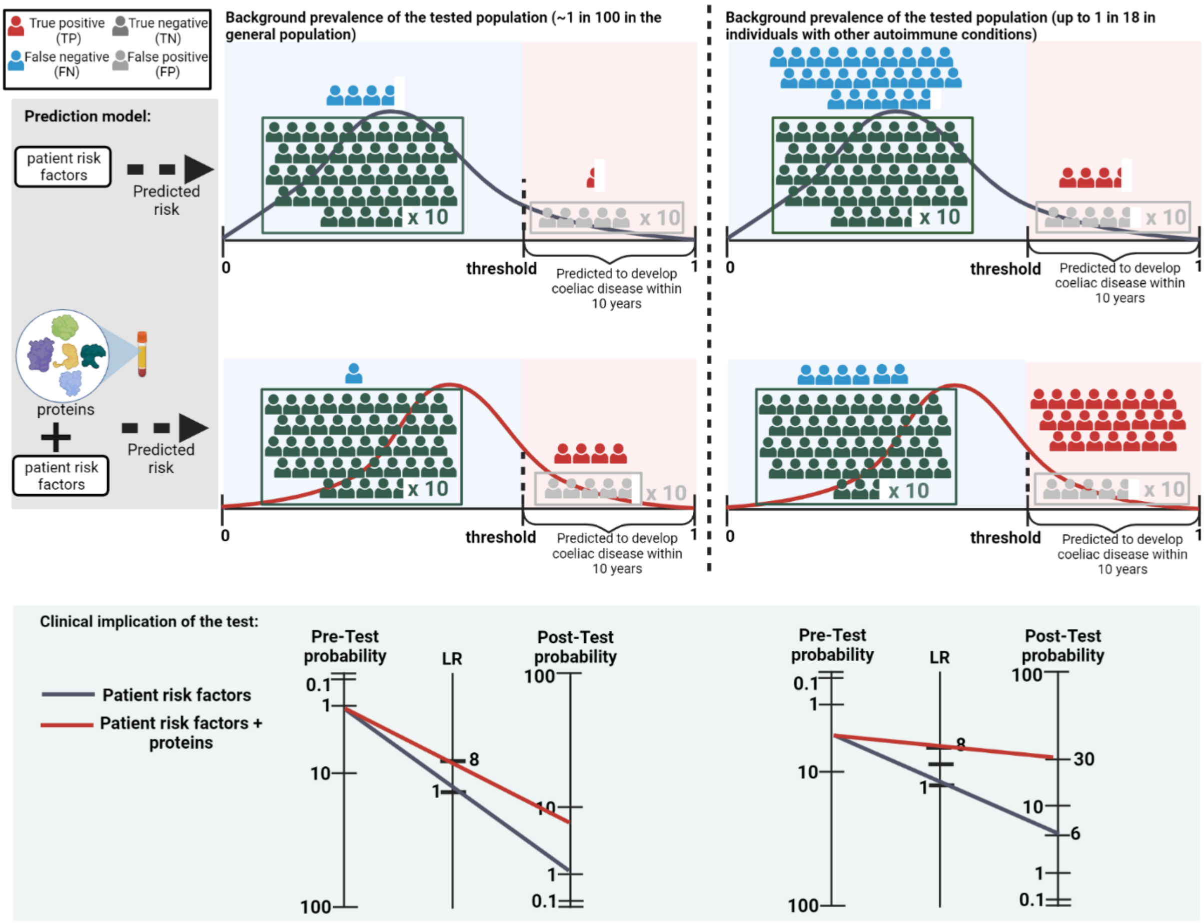

**Figure S3. Example of the improvement from proteomically informed screening strategies for coeliac disease.** We present two scenarios, in which screening is performed in 1) the general population and 2) a high-risk population (individuals with other autoimmune conditions). According to their predicted risk, individuals are classified as "positive" (those predicted to develop coeliac disease within the next 10 years) or "negatives" (not predicted at risk of coeliac disease). We illustrate the number of true positives, false positives, true negative and false negative that would be obtained according to the detection rate we estimated for coeliac disease in UK biobank at a 10% false positive rate. We further represent the pre-test probability, likelihood ratio (LR) and post-test probability in the two different scenarios (general population and high-risk population).

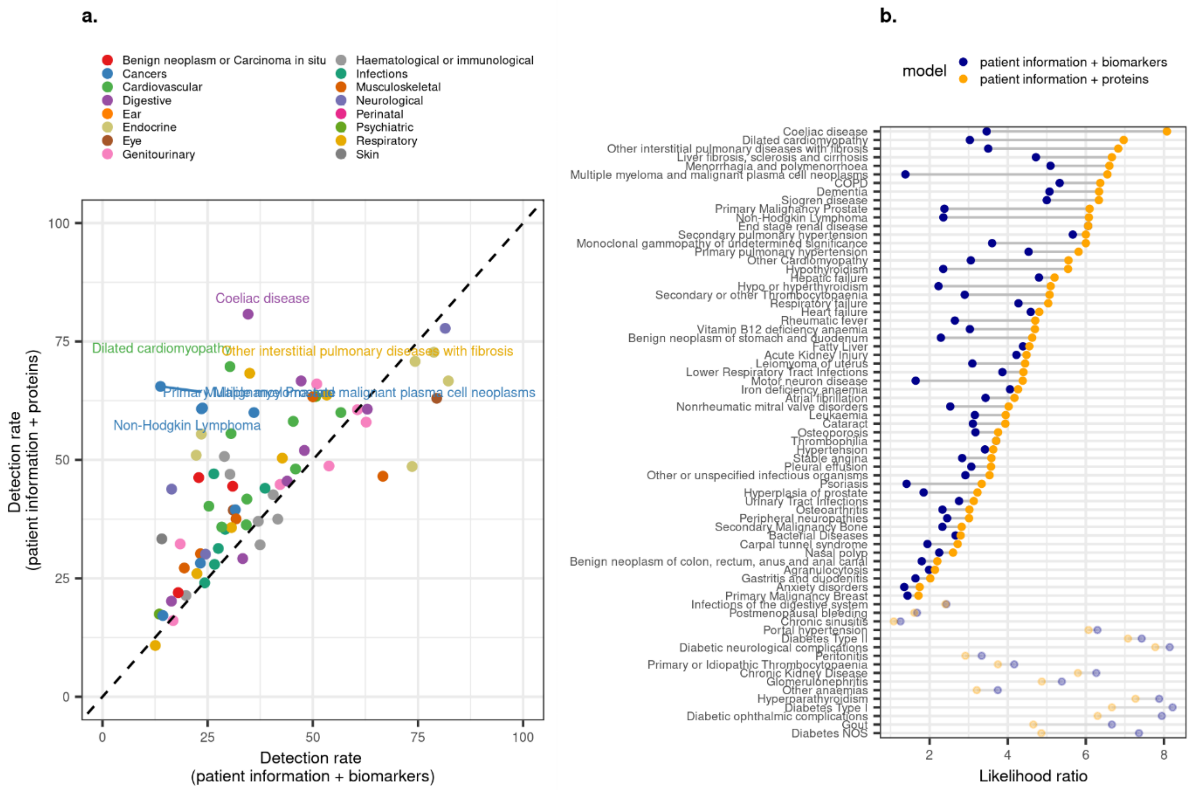

**Figure S4. Comparison of predictive performance between proteins-based (patient information + proteins) and biomarker-based (patient information + biomarkers) models. a,** Comparison between detection rates (at a 10% false positive rate) achieved by protein-based and biomarker-based models. **b,** Comparison in likelihood ratios for protein-based (orange) or biomarker-based models (grey).

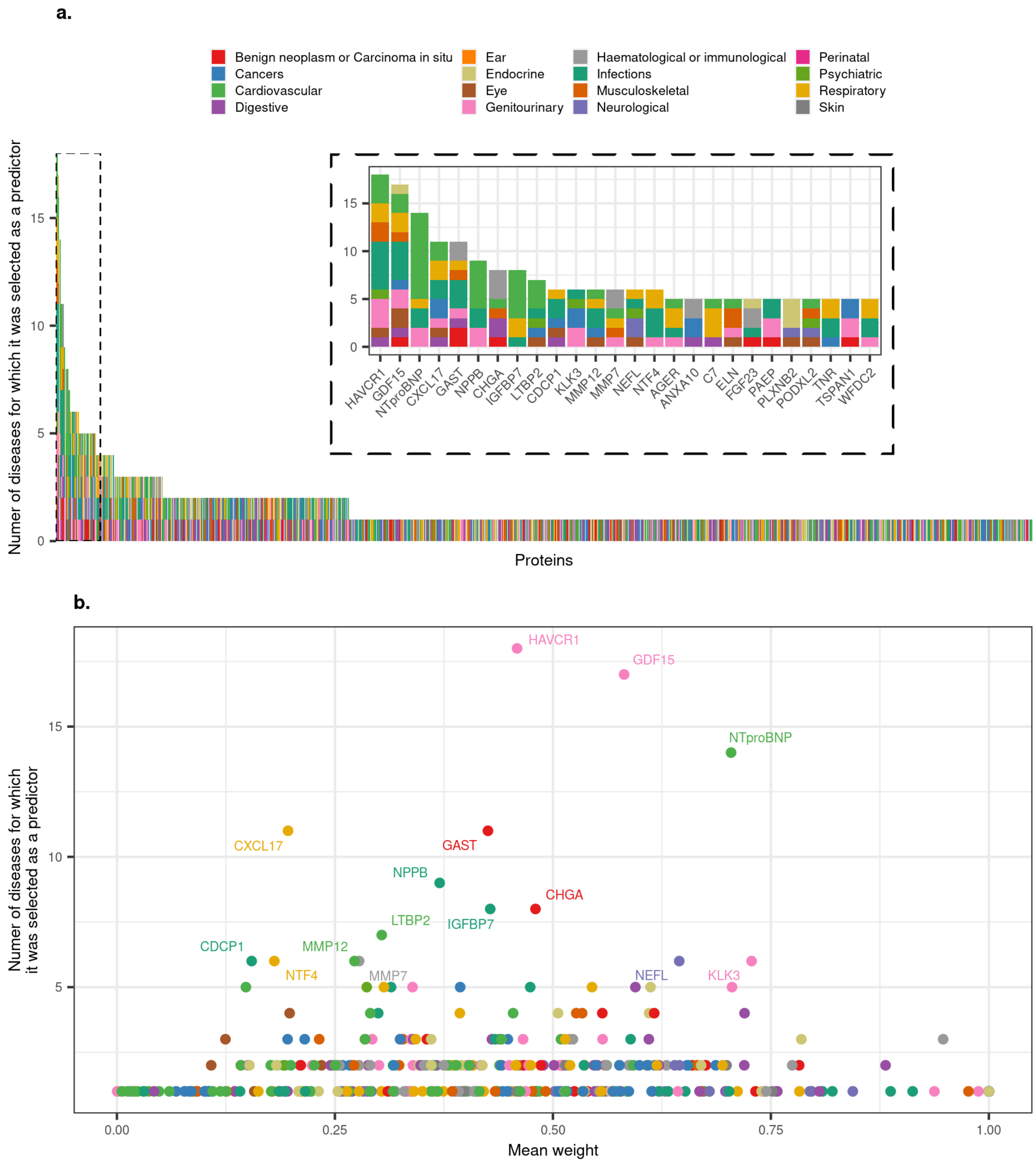

**Figure S5. Disease specificity of predictor proteins. a,** Number of individuals diseases for which a protein was selected as a predictor across the 88 diseases. These were diseases for which the C-index was significantly improved or improved by more than 0.4 over the patient information model. **b**, Average contribution of proteins across diseases. Average weights (normalised to the top predictor) from the optimised prediction models for each protein (across diseases for which it was selected as a predictor).

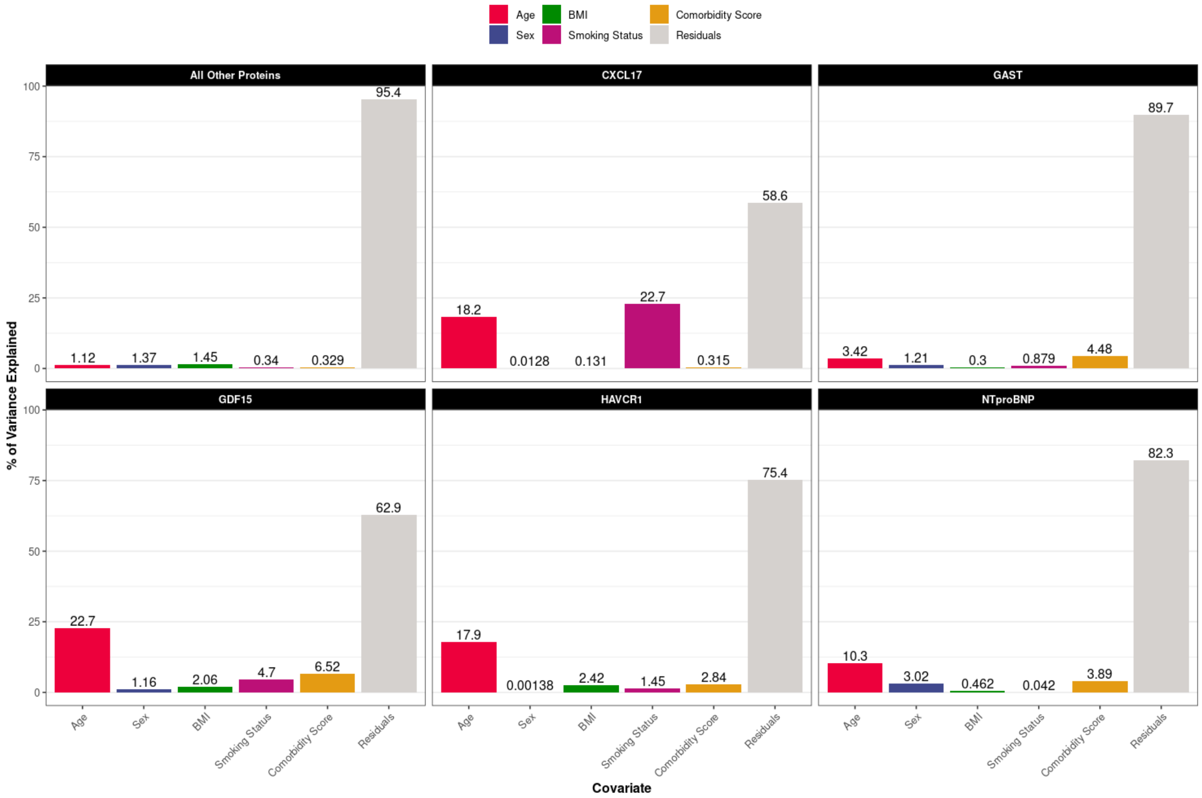

**Figure S6. Proportion of variance explained in plasma levels of proteins predictive across more than 10 diseases by demographic characteristics.** Proportion of variance by age, sex, body mass index (BMI), smoking status and a comorbidity score (see Methods) in a joint model. This is compared the average variance explained by each of these characteristics in plasma levels of all other proteins.

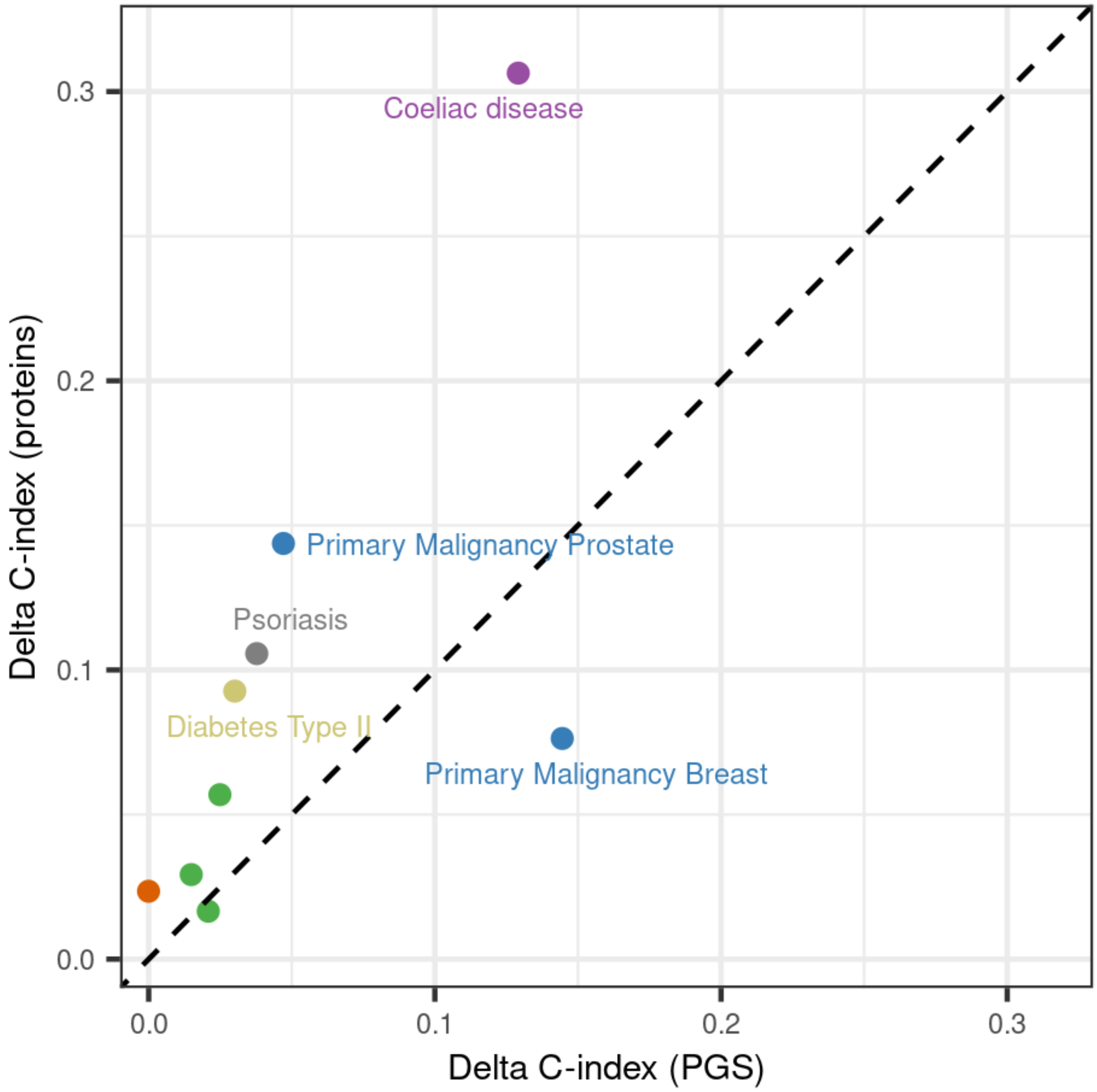

**Figure S7. Comparison of the predictive performance of protein-based and PGS over clinical models.** Comparison of the improvement in predictive performance over patient information models (delta C-index) provided by PGS and 5-20 proteins. Only 9 diseases for which either proteins or PGS provided a significant improvement in performance are shown.

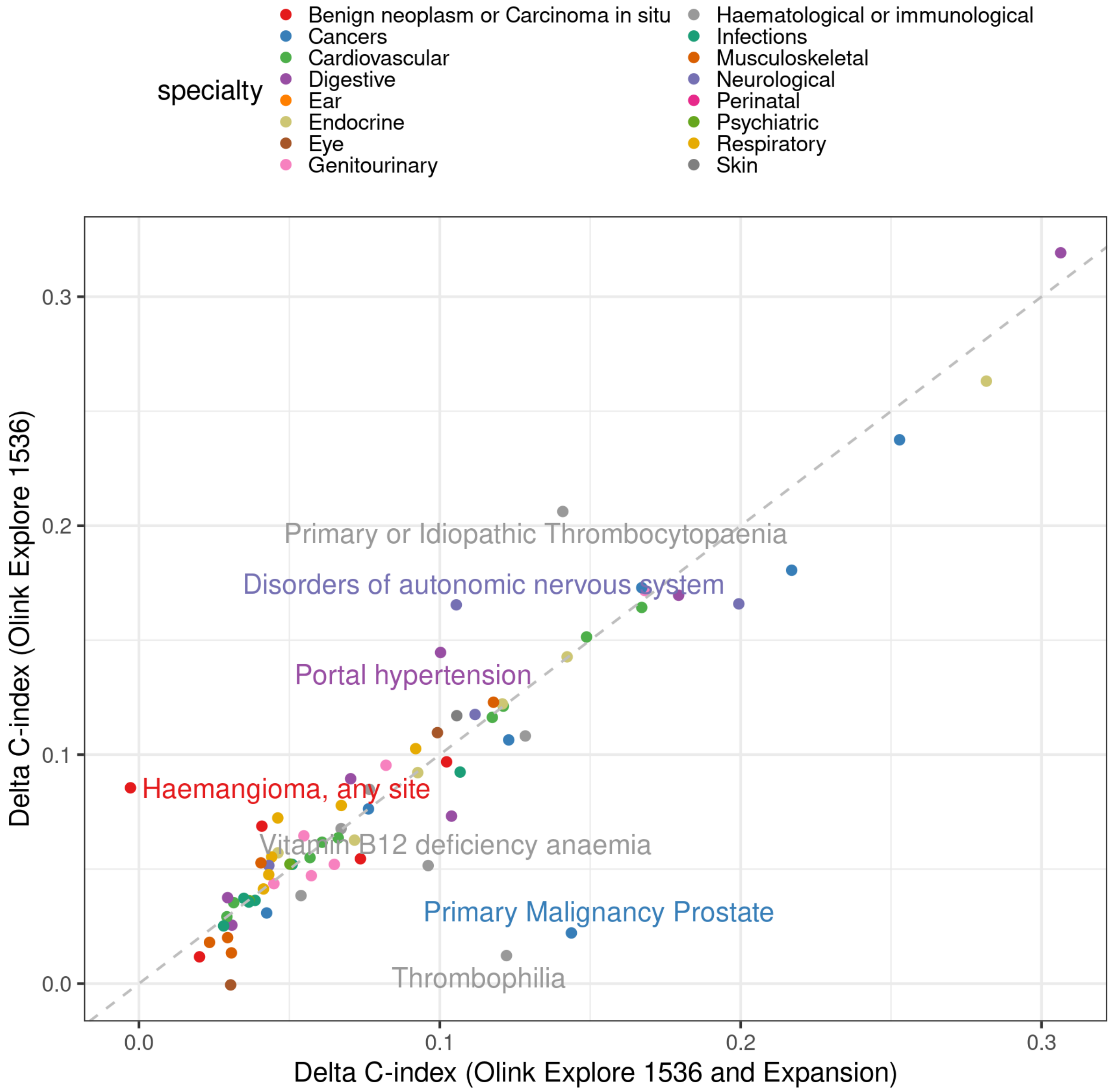

**Figure S8. Comparison of improvement in predictive performance provided from proteins derived from the Explore 1536 + Expansion and Explore 1536 platforms.** Comparison is shown for those diseases which were significantly improved by proteins for either analysis.

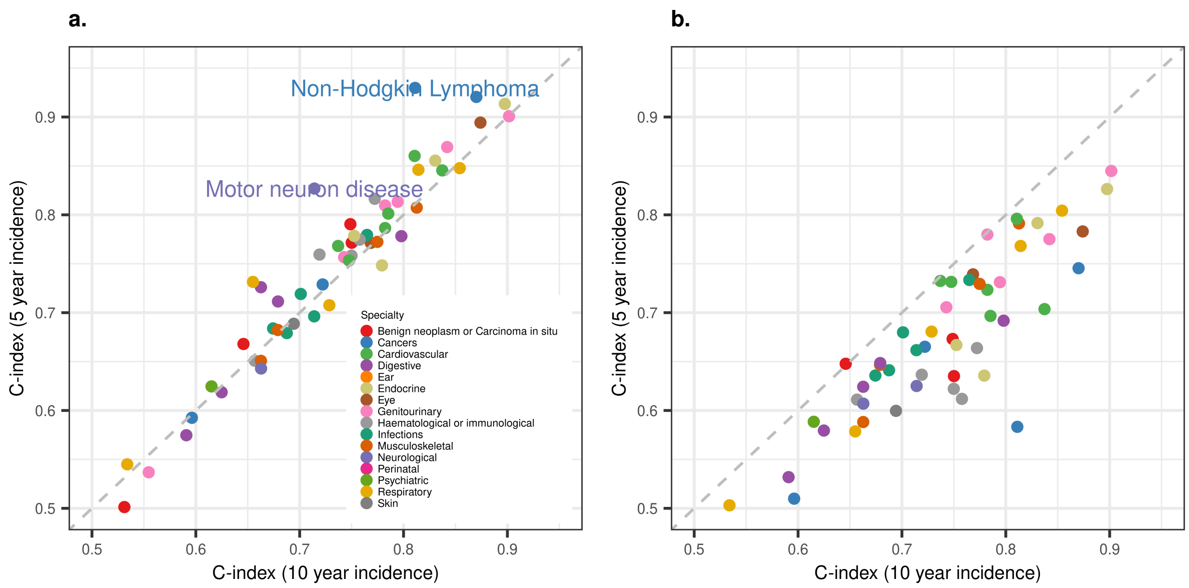

**Figure S9. Predictive performance of 10 year incidence models for 5 year incidence. a,** Protein-based models trained for prediction of 10 year incidence were tested for 5 year incidence prediction. This analysis was restricted to diseases which were improved by proteins over the patient risk factors (or improved the C-index by more than 5%), and had at least 20 incident cases during 5 years of follow-up in the test set (54 diseases). **b,** Patient information (age, sex, BMI, self-reported ethnicity, smoking status, alcohol consumption and paternal of maternal history of disease) models trained for prediction of 10 year incidence were tested for 5 year incidence prediction for the same 54 diseases. c, Improvement

**Supplementary references**

1. Sun BB, Chiou J, Traylor M, et al. Genetic regulation of the human plasma proteome in 54,306 UK Biobank participants. bioRxiv 2022:2022.06.17.496443.
2. Wik L, Nordberg N, Broberg J, et al. Proximity Extension Assay in Combination with Next-Generation Sequencing for High-throughput Proteome-wide Analysis. Mol Cell Proteomics 2021;20:100168.
3. Zhong W, Edfors F, Gummesson A, Bergstrom G, Fagerberg L, Uhlen M. Next generation plasma proteome profiling to monitor health and disease. Nat Commun 2021;12:2493.
4. Sinnott-Armstrong N, Tanigawa Y, Amar D, et al. Genetics of 35 blood and urine biomarkers in the UK Biobank. Nat Genet 2021;53:185-94.
5. Vuckovic D, Bao EL, Akbari P, et al. The Polygenic and Monogenic Basis of Blood Traits and Diseases. Cell 2020;182:1214-31 e11.
6. Hoffman GE, Schadt EE. variancePartition: interpreting drivers of variation in complex gene expression studies. BMC Bioinformatics 2016;17:483.
7. Elixhauser A, Steiner C, Harris DR, Coffey RM. Comorbidity measures for use with administrative data. Med Care 1998;36:8-27.
